# Broader functional and memory antibody responses in adult females than males in response to seasonal H3N2 vaccine virus

**DOI:** 10.64898/2026.11.17.26368332

**Authors:** Han-Sol Park, Anna Yin, Maclaine A. Parish, Jingping Hu, Ainslee Zou, Rishi Pandey, Katherine Z.J. Fenstermacher, Richard E. Rothman, Andrew Pekosz, Franklin R. Toapanta, Sabra L. Klein

## Abstract

Females of reproductive ages often mount greater antibody responses than males following influenza vaccination, but the breadth and depth of these differences, including antibody-dependent Fc receptor (FcR)-mediated functions have not been explored. Plasma samples were collected from male and female healthcare workers (range of 21-48y; mean of 31y) at 0, 28, and 180 days post-vaccination (DPV) during the 2022-2023 influenza vaccine season to assess antibody responses to A/H1N1, A/H3N2, and B/Victoria vaccine viruses. Females had greater A/H3N2, but not A/H1N1 or B/Victoria, neutralizing antibody (nAb) responses compared to males through 180 DPV, with differences being most pronounced among A/H3N2 responders (≥4 fold-change in nAb titers). Memory B cells isolated from female responders secreted more IgG in response to *ex vivo* H3 stimulation than those from male responders at 28 and 180 DPV. Before vaccination, female responders had greater anti-H3 IgG responses and FcγRIIB (CD32b) antibody binding than males, with females maintaining greater responses through 180 DPV. Female responders had greater H3-specific antibody-dependent monocyte phagocytosis (ADMP) and natural killer cell (ADNK) secretion of IFNγ and CCL4 prior to and after vaccination, with ADMP positively correlating with greater H3N2 neutralization through 180 DPV in females only. Among responders, the durability of ADMP, ADNK, and nAb in females as compared to males defined the sex differences in influenza-vaccine induced immunity. The greater FcγR-binding antibodies prior to vaccination in females might improve the durability of both neutralizing and non-neutralizing antibody responses to diverse vaccines, including next generation influenza vaccines.

**Graphical abstract:** 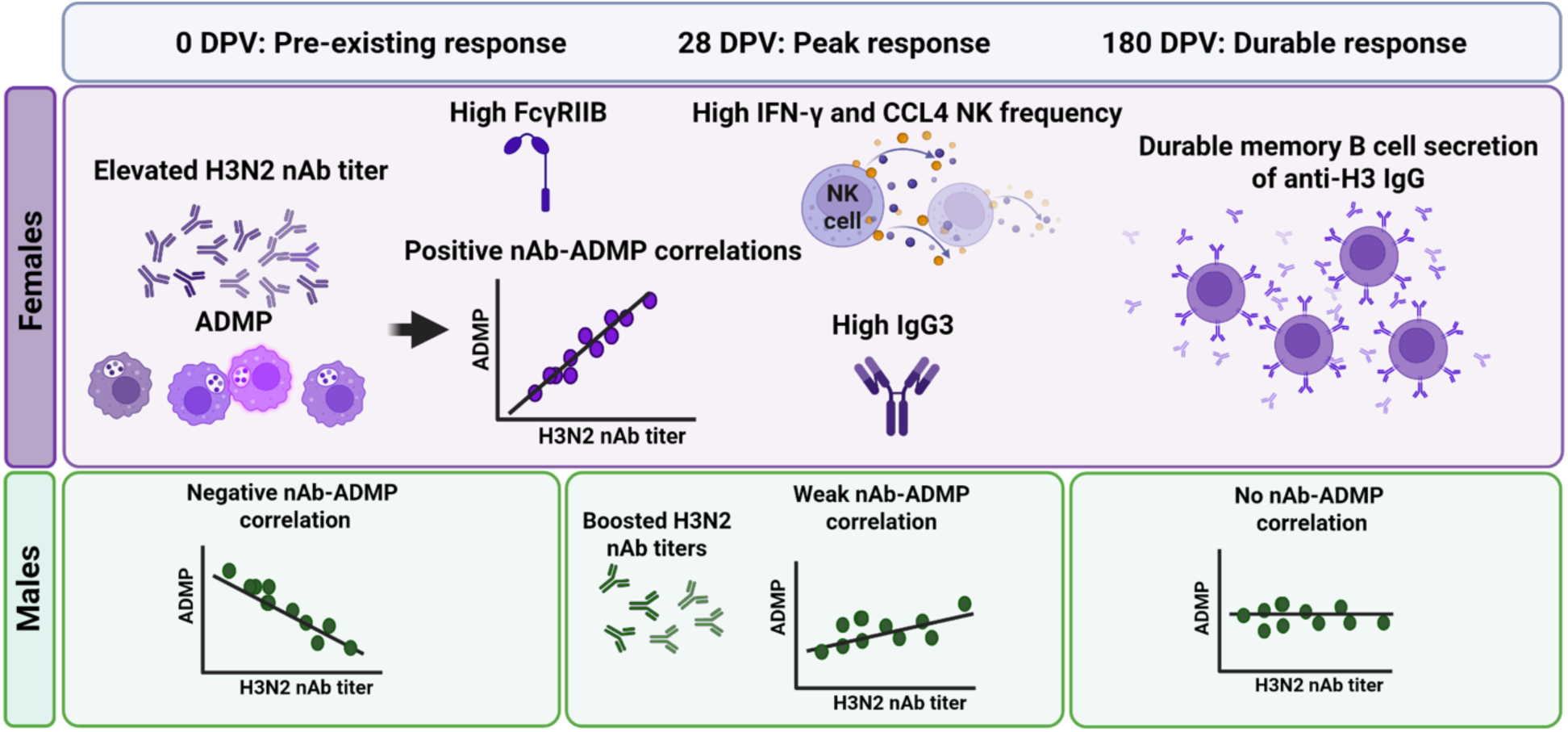

## Introduction

Biological sex (i.e., physiological differences between males and females due to sex chromosome complement, concentrations of gonadal steroids, or both) can contribute significantly to variability in influenza vaccine-induced immunity^1,2^. Of the few human influenza vaccine cohort studies that have considered sex differences in vaccine outcomes, in adults of reproductive ages (around 18-49 years), females have greater antibody titers (i.e., either neutralizing antibody [nAb] or hemagglutination inhibition [HAI] titers) than males following receipt of either seasonal or pandemic influenza vaccines^3–5^. The sex difference in antibody responses is significantly influenced by levels of pre-existing immunity to selected influenza A virus (IAV) and influenza B virus (IBV) strains, in which greater levels of pre-existing immunity mask sex differences in post-vaccine antibody titers and seroconversion^6^. An analysis of influenza vaccine effectiveness using data across seven influenza seasons reveals that vaccine effectiveness is generally greater for females than males, with this difference being greatest for A/H3N2 and B/Victoria over A/H1N1pandemic 2009^7^. Even prior to the 2009 H1N1 pandemic, adult females produced greater HAI titers against the A/H3N2 and B/Yamagata vaccine virus strains, but not against the A/H1N1 vaccine viruses^5^. During the 2017-2018 and 2019-2020 influenza seasons, adult females mounted more robust neutralizing antibody responses to H3N2 vaccine virus compared to males^6,8^.

Single cell RNA sequencing highlights that CD19^+^ B cells from reproductive age females have greater transcriptional activity than do B cells from males within one week after receipt the seasonal influenza vaccine, with upregulation of genes associated with estrogen signaling and genes with estrogen-response elements (EREs) along NF-κB-mediated TNF signaling pathways in naïve, memory, and antibody secreting B cell subsets^6^. *Ex vivo* estradiol treatment of peripheral blood mononuclear cells (PBMCs) from females, but not males, increases the number of HA^+^IgG^+^ cells, the size of individual ELISPOT colonies and the expression of ERE genes along the NF-κB-mediated TNF signaling pathway, which is inhibited by pre-treatment of cells with an estrogen receptor antagonist. A meta-analysis of 19 randomized controlled trials of influenza vaccination from 2010-2018 reveals a slightly higher rate of seroconversion and significantly greater geometric mean antibody titers in females compared to males^9^.

Sex differences, particularly in response to H3N2 vaccine viruses, are consistently observed in the antigen binding fragment (Fab)-mediated antibody functions (e.g., virus neutralization) among reproductive aged adults^4–6,8^. Whether sex differences are observed in the fragment crystallizable (Fc)-mediated antibody functions following influenza vaccination has not been explored and formed the basis of this study. Beyond neutralization, IgG antibodies engage Fcγ receptors (FcγRs) on natural killer (NK) cells, monocytes, macrophages, neutrophils, and the complement complex to mediate antibody-dependent NK cell activation (ADNK), phagocytosis by monocytes and macrophages (ADMP) and neutrophils (ADNP), and complement deposition (ADCD)^10^. Interactions of Fc with the neonatal Fc receptor (FcRn) affect IgG half-life and recycling but also play roles in immune cells by altering phagocytosis and antigen presentation^11^.

Non-neutralizing antibody functions have been linked to protection from influenza independent of neutralizing antibody titer. For example, broadly reactive antibodies with Fc-mediated functions, including antibody-dependent cellular phagocytosis (ADCP), ADNP, and ADCD have been implicated in protection from influenza virus infection^10^. While the head domain of the influenza virus hemagglutinin (HA) is the target of nAbs, the HA stalk-specific antibodies interact with FcγRs on innate immune cells to mediate protection, forming the basis for several universal influenza vaccine candidates^10,12–19^. Antibodies recognizing influenza HAs that bind to FcγRs are prevalent in the population, increase with age, and show some evidence of greater durability than nAbs^10,14,20^. For example, ADMP activity in human plasma correlates with HAI titer against both circulating and historic IAVs and remains detectable at plasma dilutions where neutralization is no longer measurable^10^. Antibody Fc glycosylation further tunes these activities to enhance or reduce ADMP^10^.

Sex differences in FcR-mediated modulation of innate immune responses following seasonal influenza vaccines are not considered. In response to a parasitic vaccine, however, females have greater frequencies of the high affinity FcγRI (CD64) on monocytes, which increases antigen presentation following vaccination in female compared with male mice^21^. In a simian immunodeficiency virus vaccine trial, female macaques have greater protection than males following virus challenge, a difference attributed in part to elevated antibody-dependent, complement-mediated lytic activity and differential antibody glycosylation and Fc-effector function between the sexes^22^. While there are many known sex differences in innate immune cell functions^23^, the extent to which vaccine-induced antibodies differentially induce innate immune cell functions in humans has not been systematically considered and formed a basis of the current study.

## Results

### Participant Demographics

Reproductive-age healthcare workers (18-49 years old) from the Johns Hopkins Health System (JHHS) were recruited during the 2022-2023 influenza vaccine season (September through October). One hundred participants (n=50/sex) were enrolled in this study prior to the receipt of the quadrivalent influenza vaccine (QIV), with ages ranging from 21 through 48 and an overall average of 31.2 ± 7.7 years old. The cohort comprised of 53% White, 29% Asian, 15% Black or African American, and 3% other, with 10% Hispanic or Latino based on self-reported data. 46% (n=23) of females reported birth-control use, with 47.8% (n=11) of those using oral contraceptives, 26.1% (n=6) using IUD, 13% (n=3) using arm implants, 8.7% (n=2) using vaginal ring, and 1% (n=1) using barrier methods (i.e., condoms). None of the females were pregnant or had a hysterectomy at the time of enrollment. No female had a history of Polyendocrine Metabolic Ovarian Syndrome in this study.

### *Females have greater neutralizing antibody responses and memory B cell antibody* secretion against H3N2 than males

Virus-specific nAb titers were measured against A/H3N2, A/H1N1 and B/Victoria 2022-2023 vaccine viruses using plasma collected at 0, 28, and 180 days post vaccination (DPV). At baseline, plasma was available for n=49 males and n=42 females (n=91 total; **Table 1**). Seroprotection status was determined at each vaccination timepoint, with nAb titers ≥ 1:40 considered as seroprotective. There were significantly lower pre-existing nAb titers against A/H3N2 and B/Victoria than A/H1N1 influenza at 0 DPV (**Figure 1a-c**). Females had significantly greater A/H3N2 nAb titers at 28 and 180 DPV than males (**Figure 1a**). Titers of nAb against A/H3N2 peaked at 28 DPV; the average relative change in A/H3N2 nAb titers between 28 DPV and 0 DPV was 7.77 for males and 8.51 for females (p=0.77). By 180 DPV, A/H3N2 nAb titers declined from 28 DPV for both sexes, with males experiencing a slightly greater average relative change (−0.39) than female (−0.23) participants (p=0.12).

**Figure 1.**
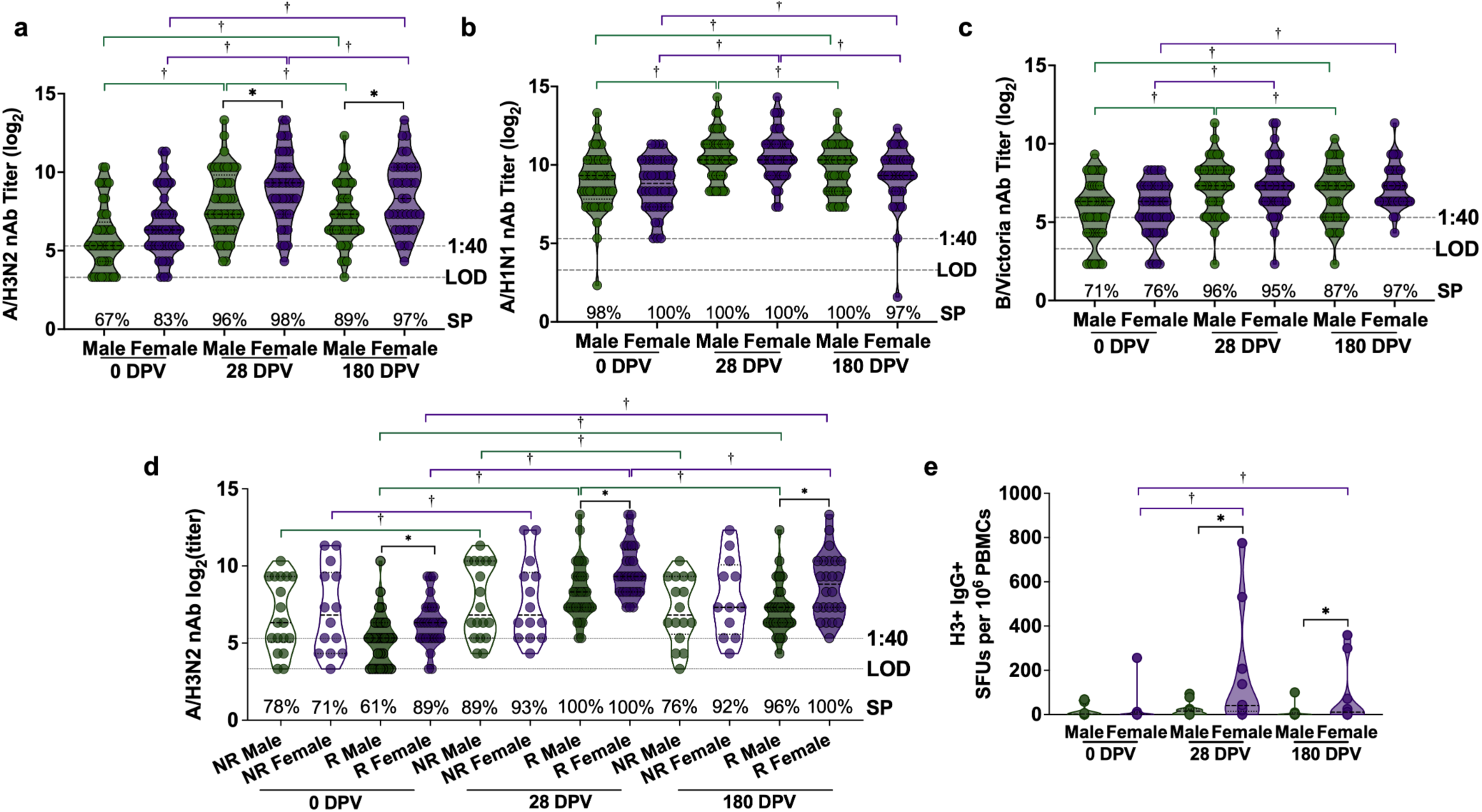
Sex differences in neutralizing and memory B cell antibody responses against H3N2 vaccine viruses are driven by responders, with females maintaining greater responses before and after vaccination. (a-c) Neutralizing antibody (nAb) titers against 2022-2023 vaccine A/H3N2, A/H1N1, and B/Victoria viruses were measured at 0, 28, and 180 days post vaccination (DPV) in plasma from n=49 males and n=42 females. **(d)** H3N2 nAb titers were analyzed by sex and H3N2 responder status (non-responder [NR] or responder [R]), defined as ≥ 4-fold change in nAb titers between 0 and 28 DPV, for n=18 NR males, n=31 R males, n=14 NR females, and n=28 R females. **(e)** H3-specific IgG^+^ spot-forming units (SFU) per 1×10^6^ PBMCs were analyzed in memory B cells by ELISpot across 0, 28, and 180 DPV in H3N2 responders by sex (n=8-11/sex). nAb titers were log_2_-transformed and analyzed by linear mixed-effects regression analysis, with an interaction term for sex and timepoint and/or H3N2 responder status, controlling for continuous age. ELISpot count data were analyzed by Poisson mixed effects regression. * represents p<0.05 sex differences, † represents p<0.05 timepoint differences. Brackets for timepoint differences are colored by sex (green=males, purple=females).

**Table 1.** Proportions of participants that seroconverted after vaccination and were seroprotected prior to vaccination against A/H3N2, A/H1N1, and B/Victoria vaccine viruses.

|  | **Male** | **Female** | **Overall** | **P-value** |
|---|---|---|---|---|
| **Age (mean, SD)** | 31.2 (6.5) | 31.2 (8.8) | 31.2 (7.7) | 0.98 |
| **A/H3N2 Seroconversion (n, %)** |  |  |  | 0.74 |
| Non-Responder (<4 FC) | 18 (37) | 14 (33) | 32 (35) |  |
| Responder (≥ 4 FC) | 31 (63) | 28 (67) | 59 (65) |  |
| **Baseline H3N2 Seroprotection (n, %)** |  |  |  | 0.08 |
| Not Seroprotected (<1:40 titer) | 16 (33) | 7 (17) | 23 (25) |  |
| Seroprotected (≥1:40 titer) | 33 (67) | 35 (83) | 68 (75) |  |
| **A/H1N1 Seroconversion (n, %)** |  |  |  | 0.28 |
| Non-Responder (<4 FC) | 30 (61) | 21 (50) | 51 (56) |  |
| Responder (≥ 4 FC) | 19 (39) | 21 (50) | 40 (44) |  |
| **Baseline H1N1 Seroprotection (n, %)** |  |  |  | 0.54 |
| Not Seroprotected (<1:40 titer) | 1 (2) | 0 (0) | 1 (1) |  |
| Seroprotected (≥1:40 titer) | 48 (98) | 42 (100) | 90 (99) |  |
| **B/Victoria Seroconversion (n, %)** |  |  |  | 0.46 |
| Non-Responder (<4 FC) | 29 (59) | 28 (67) | 57 (63) |  |
| Responder (≥ 4 FC) | 20 (41) | 14 (33) | 34 (37) |  |
| **B/Victoria Seroprotection (n, %)** |  |  |  | 0.39 |
| Not Seroprotected (<1:40 titer) | 14 (29) | 10 (24) | 24 (26) |  |
| Seroprotected (≥1:40 titer) | 35 (71) | 32 (76) | 67 (74) |  |
| **Missing baseline nAb data** | 1 (2) | 8 (16) | 9 (9) | **0.03** |
| **Total** | 50 | 50 | 100 |  |

Regardless of sex, almost all participants (99%) had nAb titers against A/H1N1 influenza that were well above the cut off for seroprotection (≥1:40 titer) prior to vaccination, suggesting strong pre-existing immunity against H1N1 (**Figure 1b**). Despite strong pre-existing nAb titers against A/H1N1 prior to vaccination, males and females similarly increased nAb titers against A/H1N1 between 0 DPV and 28 DPV with no statistically significant sex differences observed. Between 0 and 28 DPV, the average relative change in A/H1N1 nAb titers were 7.76 and 5.73 for males and females (p=0.62), respectively. By 180 DPV, nAb titers against H1N1 declined by an average relative change of −0.39 in males and −0.40 in females (p=0.90), but remained above the cut-off for seroprotection (**Figure 1b**). For B/Victoria, 74% overall seroprotection was observed prior to vaccination, with no sex differences observed in either seroprotection or seroconversion (**Figure 1c**). nAb titers against B/Victoria increased in males and females between 0 DPV and 28 DPV with average 5.79 and 4.31 relative change (p=0.65), respectively. By 180 DPV, in males there was a decline (average relative change of −0.23) that was not observed among females (average relative change of −0.05, p=0.09; **Figure 1c**).

When participants were separated based on seroconversion status into non-responders (<4 fold-change in titer from 0-28 DPV) and responders (≥4 fold-change in titer from 0-28 DPV), sex differences in A/H1N1 and B/Victoria nAb titers were still not observed, with males and females having similar seroprotection rates across timepoints regardless of seroconversion status (**Figure S1a-b**). For A/H3N2 nAb titers, sex differences were consistently apparent across all timepoints only for responders (**Figure 1d**). Prior to vaccination, the seroprotection rate among A/H3N2 female responders (n=25/28, 89%) was significantly greater than that of male responders (n=19/31, 61%; p=0.014). At 28 DPV, 100% of male and female responders had seroprotective levels of nAb. The average relative change in A/H3N2 nAb titers between 0 and 28 DPV was comparable between male (11.77) and female (12.43) responders (p=0.85), yet females maintained significantly greater nAb titers against A/H3N2 than males before and after vaccination (**Figure 1d**). At 180 DPV, the seroprotection rates were similar between male and female responders at 96% and 100% (p=0.35), respectively. The waning of H3N2 titers between 28 and 180 DPV, however, was more apparent among males, for both responders (average relative change of −0.49) and non-responders (average relative change of −0.24), whereas this reduction in immunity was not observed among either female responders (average relative change of −0.38) or non-responders (average relative change of 0.08; **Figure 1d**). These data highlight that sex differences among A/H3N2 responders across vaccination timepoints are driven by male-specific waning and female-specific durability of functional vaccine-induced nAb against A/H3N2 6 months after vaccination. We focused our subsequent functional, systems serology, and non-neutralizing assays within A/H3N2 responder participants (n=31 males and n=28 females; **Table 1**) consistent with previous published studies^24^, to further interrogate the sex differences in vaccine-induced immunity.

Because A/H3N2 responder females maintained greater A/H3N2 nAb titers than males through 180 DPV, we next determined if H3 specific memory B cells were present and whether responses differed between male and female responders. Following differentiation of memory B cells into antibody secreting cells, the frequency of H3-specific IgG^+^ memory B cells was greater among females than males at 28 and 180 DPV, but not prior to vaccination (**Figure 1e**). Males showed little anti-H3 IgG production before vaccination and the frequency of these cells (reported as spot forming units −SFU-per 1×10e6) did not change after vaccination. In contrast, females significantly increased the frequency of cells producing anti-H3 IgG at 28 and 180 DPV, as compared with 0 DPV (**Figure 1e**). Thus, females not only have greater frequencies of H3-specific total B cells that secrete IgG^6^, but also have greater frequencies of H3-specific IgG memory B cells and females maintain this immunity through at least 180 DPV.

### *The sexes differ in antibody isotypes, class switching, and binding to FcRs in response* to H3 antigen

To assess antigen-specific immunoglobulin isotype responses, IgG and IgG1-4 (**Figures 2a-e**), IgM (**Figures 2f)**, and IgA1-2 (**Figures 2g-h**) binding to 2022-2023 A/H3N2 vaccine strain HA (H3) was multiplexed in plasma from the A/H3N2 responders. Prior to vaccination, females had greater anti-H3 IgG responses than males **(Figure 2a).** After vaccination, anti-H3 IgG, IgG1-3, IgA1, and to a lesser extent IgM, IgG4, and IgA2, increased in both males and females, with peak responses occurring at 28 DPV (**Figures 2a-e**). Females had greater peak anti-H3 IgG3 than males at 28 DPV (**Figure 2d**).

**Figure 2.**
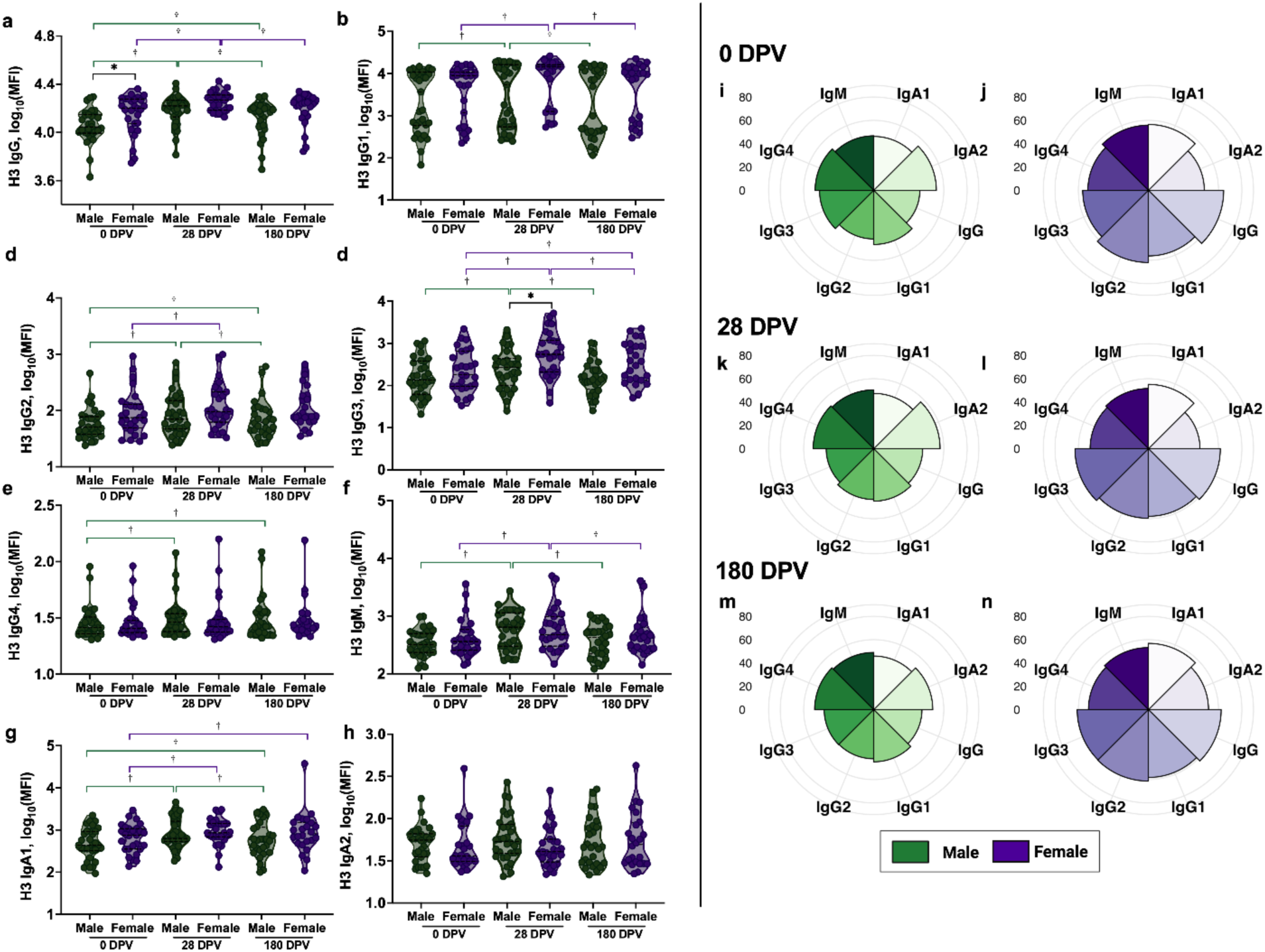
Among H3N2 responders, females have greater H3-specific IgG and IgG3 responses before and after vaccination than males. (**a-h**) H3-specific immunoglobulins were measured by systems serology. Anti-H3 immunoglobulin Mean fluorescence intensity (MFI) peaked at 28 and declined by 180 DPV. (**i-n**) Average percentile ranks of each H3-specific immunoglobulin were calculated at 0, 28, and 180 DPV and visualized by sex using polar plots. Systems serology data were log_10_-transformed and analyzed by linear mixed effect regression with an interaction term for sex and timepoint, adjusting for continuous age. * represents p<0.05 sex differences and † represents p<0.05 timepoint differences. Brackets for timepoint differences are colored by sex (green=males, purple=females).

Because females had greater A/H3N2 nAb titers and greater anti-H3 IgG and IgG3 responses than males prior to and after vaccination, we next compared the relative anti-H3 antibody isotype features between males and females over time using polar plots, in which each spoke represented one measured antibody feature and each value represented the median percentile rank for females compared with males at either 0, 28, or 180 DPV (**Figure 2i-n**). Prior to and more so 28 and 180 DPV, females maintained greater proportional anti-H3 IgG responses than males, with coordinated elevation of IgG and IgG1-3 in females compared with males after vaccination (**Figure 2i-n**). Among males, anti-H3 IgG4, which is less functional for Fc-mediated responses to viral antigens with greater anti-inflammatory properties^25,26^, was proportionally greater than in females 28 and 180 DPV (**Figure 2i-n**).

We next assayed the ability of H3-specific antibodies to bind FcγRIIA-R, FcγRIIA-H, FcγRIIB, FcγRIIIA-V, FcγRIIIA-F, FcγRIIIB, FcRn, and FcαR. For each assay, the mean fluorescence intensity (MFI) of FcR protein-conjugated to PE for each antigen was quantified for antigen-specific FcR binding responses (**Figure 3a-h**). Prior to vaccination, the binding of anti-H3 IgG to FcγRIIB (CD32b) was greater in females than males (**Figure 3c**). Greater pre-existing IgG binding to FcγRIIB increases nAb responses in humans and mice^27,28^. Sex differences were not observed in the binding of H3-specific antibodies to FcγRIIA-H, FcγRIIIA-V, FcγRIIIA-F, FcγRIIIB, or FcRn (**Figure 3a, d-g**). Binding of antibodies, particularly IgG to FcγRs, declined in both males and females at 180 relative to 28 DPV (**Figure 3**), mirroring the peak in antibody titers at 28 DPV (**Figure 2**). Females maintained greater proportional anti-H3 IgG binding to FcγRIIB and FcRn than males both prior to and 28 and 180 DPV (**Figure 3i-n**). In contrast, males showed evidence of greater proportional binding of anti-H3 IgG to FcγRIIA (CD32a) both prior to and 28-180 DPV (**Figure 3i-n**).

**Figure 3.**
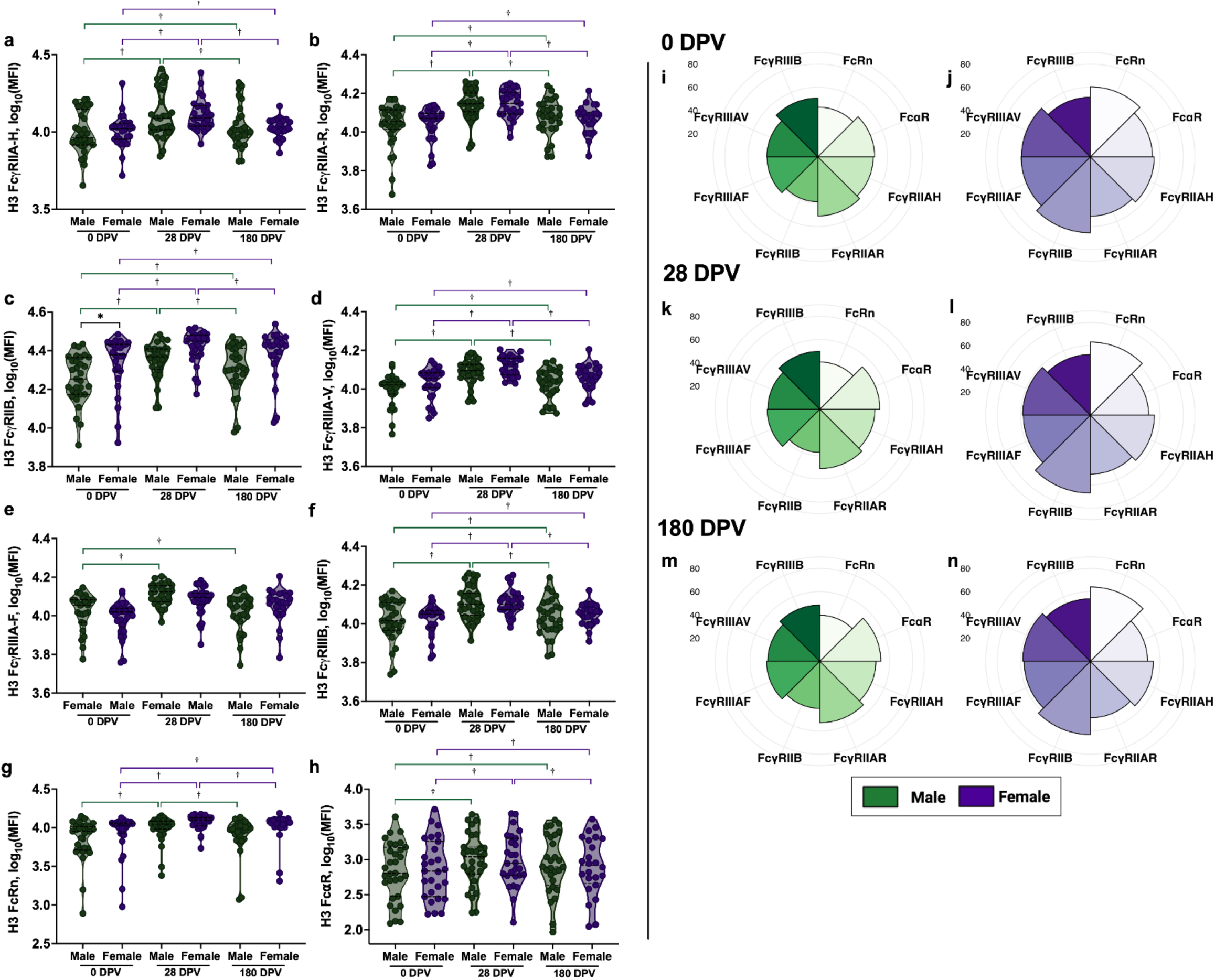
Among H3N2 responders, anti-H3 antibody binding to Fc*γ*Rs peaks at 28 DPV, with females only having greater anti-H3 antibody binding to Fc*γ*RIIB than males prior to vaccination. (**a-h**) H3-specific antibody binding to FcRs were measured by systems serology and reported as mean fluorescence intensity (MFI). (**i-n**) Average percentile ranks of each anti-H3 immunoglobulin bound to Fc*γ*Rs were calculated at 0, 28, and 180 DPV and visualized by sex using polar plots. Systems serology data were log_10_-transformed and analyzed by linear mixed effect regression with an interaction term for sex and timepoint, adjusting for continuous age. * represents p<0.05 sex differences and † represents p<0.05 timepoint differences. Brackets for timepoint differences are colored by sex (green=males, purple=females).

### *Fc*γ*R-mediated non-neutralizing antibody functions against H3 are greater in females* than males

While sex differences in H3-binding antibodies were minimal, we hypothesized that the function of H3-specific antibodies binding to FcRs might differentially affect innate immune cell activation. Antibody-dependent complement deposition was measured as C3 deposition (MFI) on H3-coupled beads opsonized with H3-specific antibodies. There was no effect of either sex or vaccination on ADCD (**Figure S2a**). Flow cytometry was used to measure ADNP, ADNK and ADMP in response to H3-specific antibodies bound to H3-coupled bead (**Figure S3**). Because CD16 (FcγRIII) is cleaved by matrix metalloproteinase and ADAM17 upon activation in NK cells^29,30^, neutrophils^31,32^, and monocytes^33^, we did not measure its expression by flow cytometry. Instead, FcγR-expressing subsets were defined using CD64 (FcγRI), CD32a (FcγRIIA), and CD32b (FcγRIIB) for monocytes, and CD64 and CD32a for neutrophils. NK cells (CD3^-^CD56^+^CD66b^-^CD14^-^) were not subdivided by FcγR expression.

Antibody-dependent neutrophil phagocytosis was measured by flow cytometry using H3-coupled beads to generate a PhagoScore^34,35^ for neutrophil activity induced by antibodies recognizing H3 among H3N2 male and female responders. Overall, H3-specific antibodies did not induce ADNP differently between males and females in either total, CD64^+^, or CD32a^+^ neutrophils (**Figure S2b-d**). There was greater total ADNP activity at 28 DPV compared to baseline among male responders (**Figure S2b**), but otherwise, ADNP activity did not change with influenza vaccination. Antibody-dependent NK cell activity was measured by flow cytometry using H3-coupled beads with analyses of degranulation (CD107a^+^), cytotoxic granule content (granzyme B^+^), cytokine (IFNγ^+^), and chemokine (CCL4^+^ or MIP^-^1β^+^) production in response to H3-specific antibodies from male or female A/H3N2 responders. At each vaccination timepoint, H3-specific antibodies from male and female A/H3N2 responders similarly induced NK-degranulation and granzyme B content (**Figure 4a-b**). At 180 DPV, male responders had greater frequencies of CD107a^+^ H3-specific NK cells than at 0 and 28 DPV, whereas female responders had lower frequencies (**Figure 4a**). Granzyme B^+^ H3-specific NK cell frequencies peaked at 28 DPV in both female and male responders (**Figure 4b**). Sex differences were observed for H3-specific antibody induced cytokine and chemokine production by NK cells across all timepoints, with H3-specific antibodies from female responders inducing greater frequencies of NK cells secreting IFNγ and CCL4 than antibodies from male responders (**Figure 4c-d**). H3-specific antibody induction of IFNγ and CCL4 positive NK cells were greater at 28 and 180 DPV relative to baseline for female responders, with these NK cell frequencies remaining similar across timepoints for males (**Figure 4c-d**). Despite both ADNK cytokine secretion and A/H3N2 nAb being greater in females than males, these responses were only weakly correlated with each other and did not differ by sex across timepoints, except for % of Granzyme B^+^ H3-specific NK cells in males at 28 DPV (r=0.57, p=0.0011) and % of CCL4^+^ H3-specific NK cells in females at 180 DPV (r=0.48, p=0.018).

**Figure 4.**
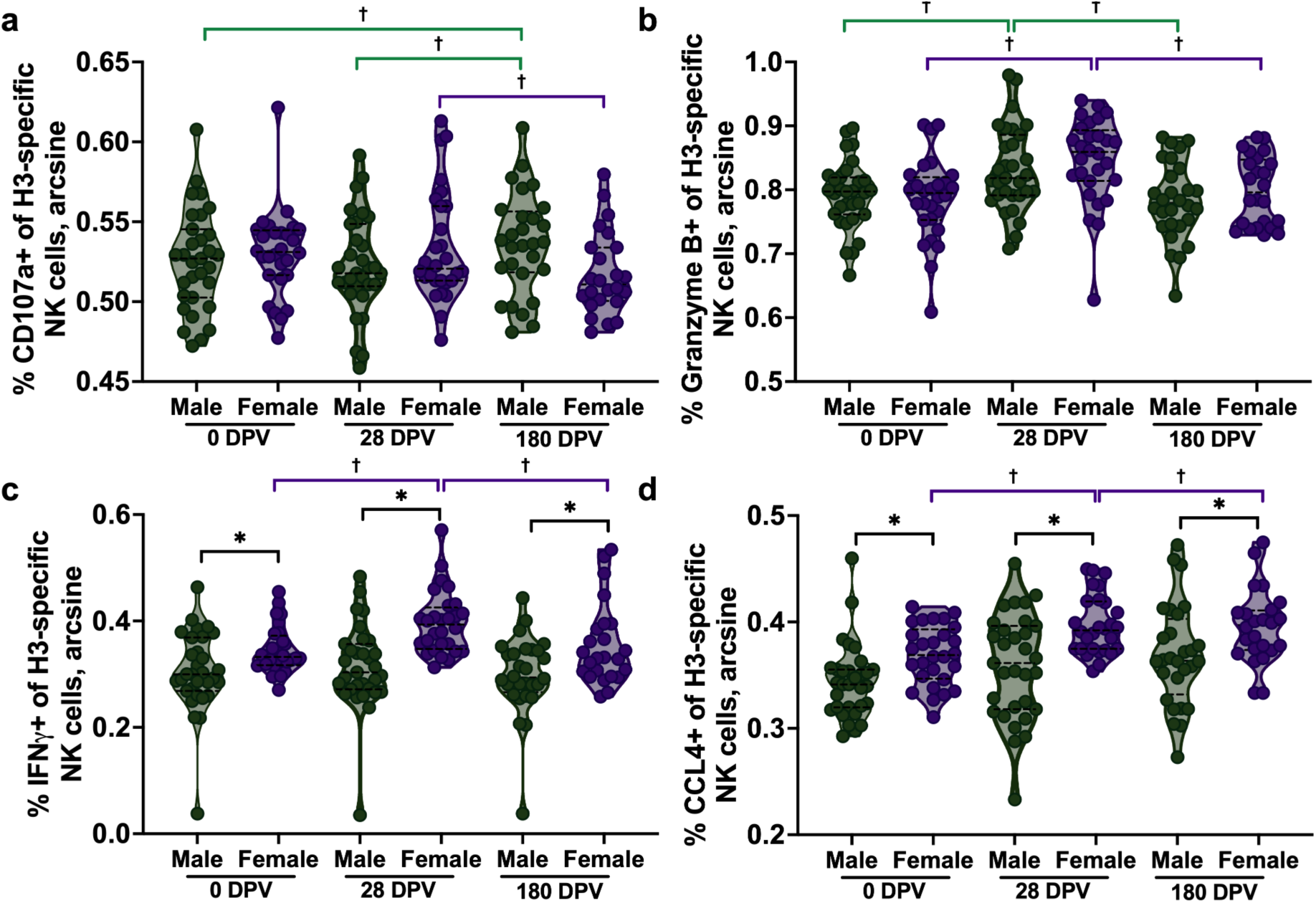
Antibody-dependent NK (ADNK) cell activity is greater among female than male H3N2 responders. Natural killer (NK) cell degranulation (**a**; CD107a^+^), cytotoxic granule content (**b**; Granzyme B^+^), cytokine (**c**; IFNγ^+^), and chemokine (**d**; CCL4^+^) functions were assessed by flow cytometry gating on H3-specific cells. Plasma from H3N2 responders collected at 0, 28, and 180 DPV was mixed with H3-coupled bead complexes and used to stimulate ADNK activity. ADNK cell frequencies were arcsine-square root transformed and analyzed with linear mixed-effects regression modeling for sex and time differences, adjusting for continuous age. * represents p<0.05 sex differences and † represents p<0.05 timepoint differences. Brackets for timepoint differences are colored by sex (green=males, purple=females).

Antibody-dependent primary monocyte phagocytosis (ADMP)^35^ was assessed by flow cytometry using H3-coupled beads to generate a PhagoScore^34,35^ for monocyte activity induced by antibodies from H3N2 responders (**Figure 5**). Across all the subsets of monocytes, females consistently entered the season with higher phagocytic activity compared to males (**Figure 5a-e**). Total, CD64^+^, CD32a^+^, and CD32b^+^ ADMP were greater in females at 0 and 28 DPV, with both males and females experiencing increases in ADMP between 0 and 28 DPV (**Figure 5a-d**). Between 28 and 180 DPV, total, CD32a^+^, and CD32b^+^ ADMP decreased in females, but ADMP levels were sustained at lower levels in males (**Figure 5a, c-d**). Compared to baseline, males had greater total, CD64^+^, CD32a^+^, and CD32b^+^ ADMP at 180 DPV, which was not observed in females, except for CD64^+^ ADMP (**Figure 5a-e**). Prior to vaccination, females had a positive correlation between total ADMP and A/H3N2 nAb titers, while males had a negative correlation (**Figure 6a**). The sex differential association between H3-specific ADMP and A/H3N2 nAb was observed across monocyte subpopulations based on FcγR expression (**Figure 6b**). At 28 DPV, both males and females had positive correlations between H3-specific ADMP and A/H3N2 nAb, with females maintaining stronger correlation than males across all monocyte subpopulations (**Figure 6c-d**). By 180 DPV, females still maintained a positive correlation between H3-specific ADMP and A/H3N2 nAb, whereas in males the correlative strength waned among total, CD64^+^, CD32a^+^, and CD32b^+^ cells (**Figure 6e-f**). Together, these data highlight that females enter the influenza season with and maintain more durable functional non-neutralizing antibody responses to H3N2 than males, which at least for ADMP is associated with greater nAb titers over 180 DPV for females only.

**Figure 5.**
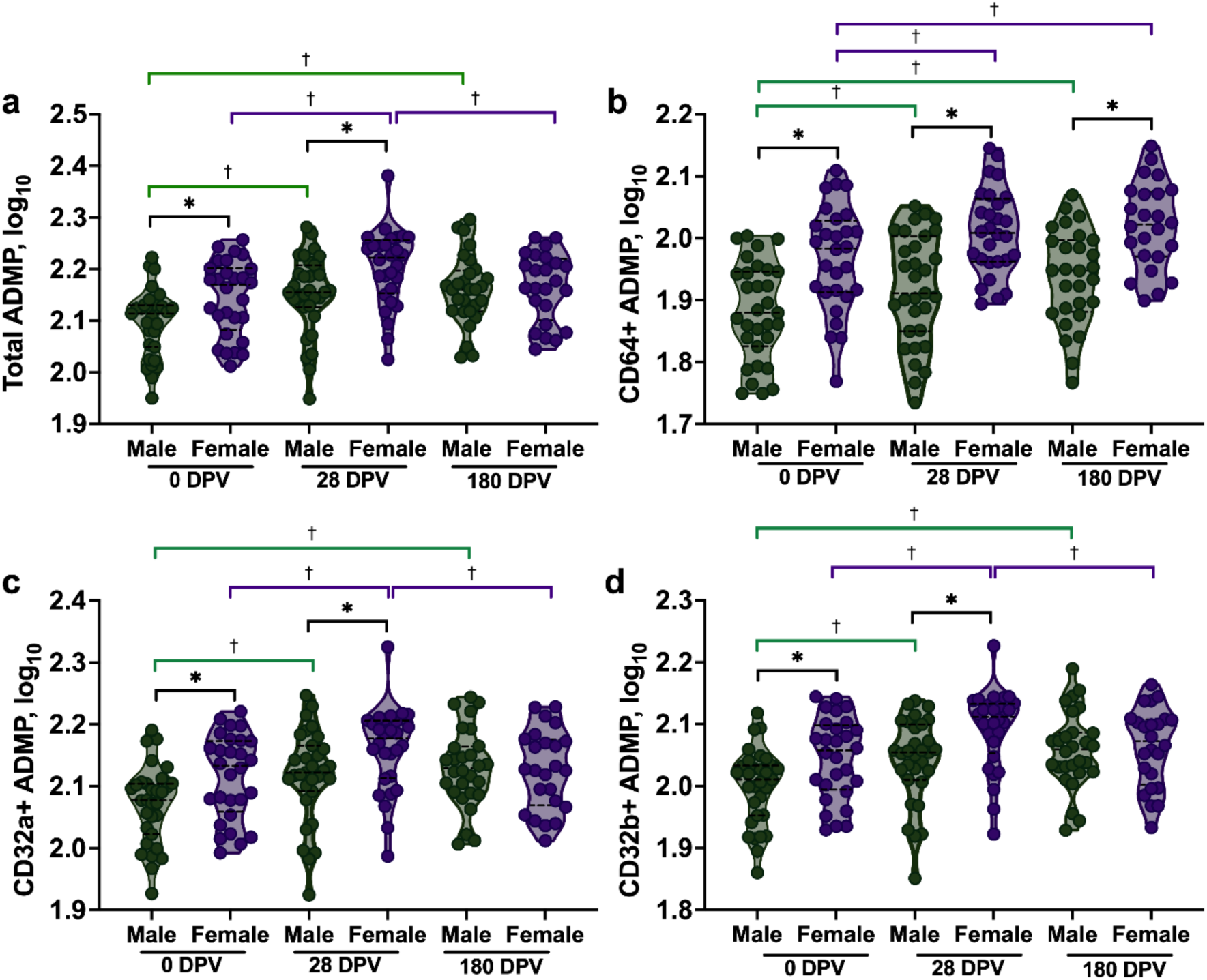
Antibodies from female H3N2 responders induce greater antibody-dependent monocyte phagocytosis (ADMP) than antibodies from males before and after vaccination. ADMP was measured by flow cytometry using plasma samples collected at 0, 28, and 180 DPV from H3N2 responders that were mixed with H3-coupled beads to form immune complexes before stimulating donor monocytes. (**a**) Total, (**b**) CD64^+^, (**c**) CD32a^+^, and (**d**) CD32b^+^ ADMP was greater in response to antibodies from H3N2 responder females than males, particularly at baseline (0 DPV) and 28 DPV. ADMP PhagoScores were log_10_-transformed and analyzed with linear mixed-effects regression modeling, adjusting for continuous age. * represents p<0.05 sex differences and † represents p<0.05 timepoint differences. Brackets for timepoint differences are colored by sex (green=males, purple=females).

**Figure 6.**
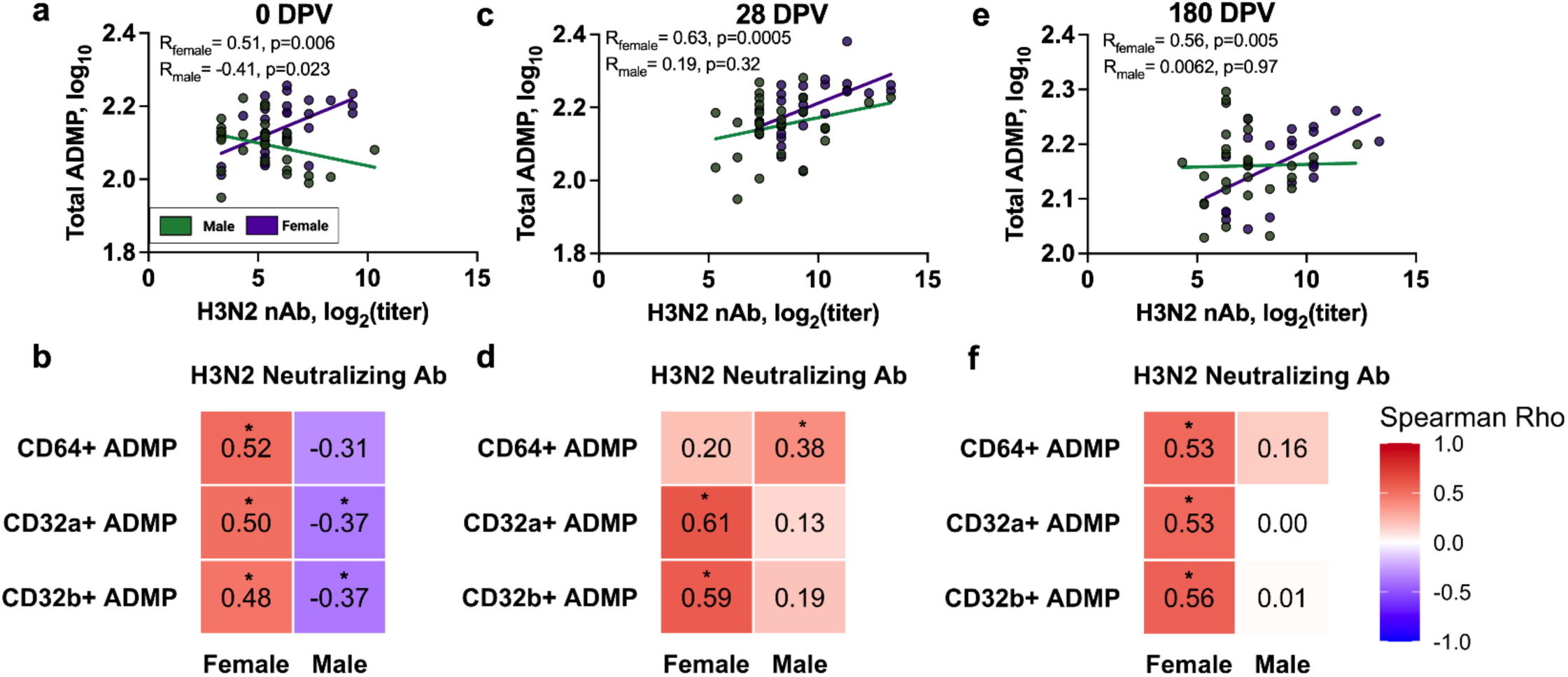
Antibodies from H3N2 responder females, but not males maintained concordant and durable ADMP and H3N2 neutralization functions before and after vaccination. Spearman correlation coefficients were calculated for total, CD64^+^, CD32a^+^, and CD32b^+^ antibody-dependent monocyte phagocytosis (ADMP) of H3-coupled beads, generating a PhagoScore using plasma samples collected at (**a-b**) 0, (**c-d**) 28, and (**e-f**) 180 DPV. PhagoScore data and H3N2 nAb were log_10_-transformed. Spearman correlation coefficients were calculated by sex at each timepoint. p<0.05, as represented by *, were considered statistically significant.

### *H3N2 ADMP, ADNK, and nAb are critical antibody features that distinguish vaccinees by* sex

Sex differences in IgG isotypes and binding to FcγRs as well as induction of non-neutralizing and neutralizing antibody responses to H3N2 were apparent in many cases through 180 DPV. Whether these responses were sufficient to distinguish the sex of the vaccinee was explored using a supervised machine learning algorithm (i.e., sparse partial least discriminant analysis [sPLS-DA]). We integrated all serological measures, including H3N2 nAb, anti-H3 isotypes, H3-specific antibody binding to FcRs, ADCD, ADNK, ADNP, and ADMP data to determine which immunological features best classified males versus females at each vaccination timepoint using complete data (n=27 females, n=31 males for 0 and 28 DPV, n=24 females, n=28 males for 180 DPV). Overall, the serological profiles of males and females were more distinguishable at 0 and 180 than at 28 DPV (**Figure 7a-c**). CD64^+^, CD32b^+^, and CD32a^+^ ADMP, CCL4^+^NK cells, IFNγ^+^ NK cells, H3N2 nAb, and anti-H3 IgG binding to FcγRIIB were among the top LASSO-selected features that best distinguished males and females, with each of these responses being greater among females than males at 0 DPV (**Figure 7a**). At 28 DPV, CCL4^+^ NK cells, IFNγ^+^ NK cells, CD64^+^ ADMP, H3N2 nAb, anti-H3 IgG3, and binding to FcγRIIB were selected as the top features separating males and females, with females having greater responses than males for all measures (**Figure 7b**). At 180 DPV, CD64^+^ ADMP, H3N2 nAb, anti-H3 IgG3, CCL4^+^ NK cells, IFNγ^+^ NK cells, anti-H3 IgG2, anti-H3 IgG binding to FcγRIIB, anti-H3 IgG, anti-H3 IgG binding to FcγRIIIA-V, and anti-H3 IgG1 were the model-selected features that distinguished sex, with females having greater responses than males for all features (**Figure 7c**). These results highlight that the greater neutralizing and non-neutralizing features of antibodies against A/H3N2, particularly at 0 and 180 DPV, separate female from male vaccinees, with all responses being upregulated in females compared to males.

**Figure 7.**
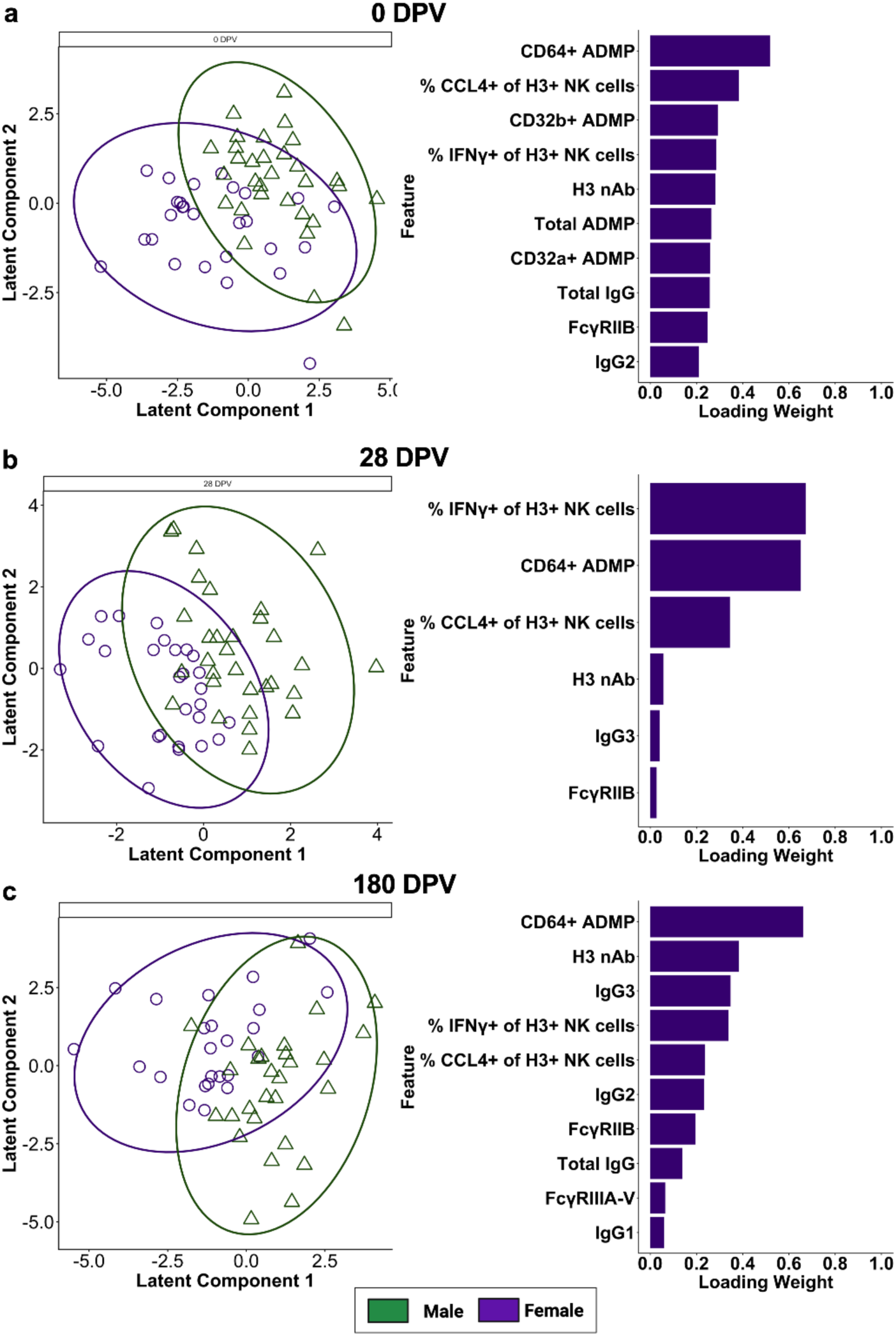
Among H3N2 responders, the serological features, including ADMP, ADNK, and nAbs are best predictors of vaccinee sex, with all responses being greater in females than males. Principal component analysis (PCA) and sparse partial least discriminant analysis (sPLS-DA) were used to identify measures that best classified or distinguished females versus males, with a model generated for (**a**) 0, (**b**) 28, and (**c**) 180 DPV. Error rates were 28.4%, 27.4%, and 31.7%, respectively. Features included for modeling were the same across timepoints, with each model using timepoint-specific data. 95% confidence ellipses for PCA are shown, colored by sex. Mahalanobis distance and 5-fold cross-validation with 50 repeats were used to tune the final model and to evaluate model performance. Loading plots show the top 10 LASSO-selected features for models with 10+ selected features. All data were z-scaled prior to modeling.

## Discussion

In the current study, while antibody responses to the A/H3N2, A/H1N1, and B/Victoria components of the seasonal 2022-2023 influenza vaccine were evaluated, only responses to A/H3N2 exhibited sex differences. In the 2022-2023 influenza season (October 2022-September 2023), the primary circulating strain was A/H3N2, with the season starting earlier than usual and cases peaking to 26.3% in December 2022 according to the U.S. Center for Disease Control (CDC)^36^, with no sex disaggregated data available. Virologic surveillance reported the majority (97.3%) of cases being IAV with A/H3N2 being the dominant strain and the remaining cases (2.7%) being IBV^36^. According to the CDC, most of the IAV and IBV that were circulating were of the same genetic clade and antigenically similar to the vaccine viruses selected for that season’s influenza vaccine^36^. For 2022-2023, the vaccine A(H1N1)pdm09-like virus had not changed from the prior year, whereas the Darwin/9/2021 (H3N2)–like virus and B/Victoria/Austria/1359417/2021–like virus were updated for egg-based vaccines in this season^37^. With the increased mutation rates in H3N2 relative to either H1N1 or IBVs, baseline immunity tends to be lower and less likely to mask sex differences^38^. In our study, the overall seroprotection rates were high for males and females alike, with 60-100% of participants having seroprotective antibody levels against A/H1N1>B/Victoria> A/H3N2 prior to vaccination. When participants were analyzed based on the seroconversion status between 0 and 28 DPV, the seroconversion rate was the greatest for A/H3N2, with 63% and 67% of males and females considered as responders (**Table 1**). Seroconversion rates for A/H1N1 and B/Victoria were lower at 44% (39% of males and 50% of females) and 37% (41% of males and 33% of females), respectively, with no sex differences observed.

Sex differences in anti-H3 IgG class switching and the binding of H3-specific antibodies to FcRs were isotype and FcR specific. Whereas females had greater anti-H3 total IgG before vaccination and IgG3 after vaccination, males had proportionally greater anti-H3 IgG4 after vaccination. IgG3 can enhance virus neutralizing antibodies as well as antibody-dependent cellular cytotoxicity and phagocytosis, which in the case of human immunodeficiency virus, promotes polyfunctional broadly neutralizing antibody responses^39,40^. In contrast, IgG4 has anti-inflammatory properties that are associated with reduced virus neutralization and activation of FcR-mediated effector functions, with proportional increases in IgG4 being associated with reduced vaccine-induced immunity^26^. Sex differences in the roles of IgG3 versus IgG4 in the broader functional attributes of antibodies have not been considered previously. Systemic lupus erythematosus (SLE) is an autoimmune disease caused by antibodies recognizing self, double stranded DNA that occurs at a ratio of 9:1 in females compared with males^41^. Serum IgG3 is elevated during active SLE disease and reduced when patients go into remission^42^. In contrast, IgG4 autoantibodies reduce SLE progression in patients, in part by reducing complement activity^39,40,43^. Together, these results highlight that the sex differences in IgG class switched responses likely impact both immunity to foreign vaccine antigens and self-antigens.

Sex differences in antibody binding to FcRs have not been reported. We observed that in response to H3 antigen, females had greater proportional binding to both FcRn and FcγRIIB. Moreover, anti-H3 IgG binding to FcγRIIB were predictive of immunity in vaccinated female participants, both prior to and 180 DPV. Previous studies highlight that pre-existing antibodies that bind FcγRIIB are associated with greater influenza virus neutralization, in part through virus opsonization^24^. While antibody binding to FcRn was not a key feature for deciphering influenza immunity between the sexes, females maintained greater proportional antibody binding to FcRn than males prior to and after vaccination. FcRn plays critical roles in antibody recycling and transport across respiratory epithelial cells^44,45^ and binding of antibody to FcRn is associated with protection against influenza^46^.

The magnitude and durability of neutralizing and non-neutralizing antibody responses against A/H3N2 were consistently greater in female than male responders and were among the most important features differentiating immunity in females compared with males. Female responders had greater nAb titers against A/H3N2 and a higher frequency of anti-H3 IgG memory B cells than males both 28 and 180 DPV highlighting that traditional correlates of protection against influenza were greater for reproductive aged females than males, as reported previously^6^. In addition to the traditional correlates of protection, H3-specific antibodies from females were better able to induce NK cell secretion of IFNγ and CCL4 both prior to and after vaccination, with responses in females peaking at 28 DPV. In contrast, there were no changes over time in H3 antibody-induced cytokine secretion by NK cells in males. H3-specific antibodies from females also were better at inducing ADMP both prior to and after vaccination including subpopulations of monocytes expressing either FcγRI or FcγRIIA/B. The durability of the sex differences was greatest among monocytes expressing the high affinity FcγRI. Analyses of Fc-mediated antibody functions provide a more in depth analysis of the vaccine-induced antibodies that may be contributing to protective effects^14^. Our data highlight that broader assessment of vaccine induced antibodies and consideration of sex should be *a priori* parameters used to analyze influenza vaccine studies.

This study was conducted among healthcare workers, which may not reflect the patterns of immunity in the general population due to work-related requirements for annual vaccination. Once we limited analyses to the H3N2 responder males and females, sample sizes were reduced, limiting the robustness of our analyses; despite this, our analyses replicated sex differences in nAb titers from 2017-2018^10^ and 2019-2020^6^ cohorts. Finally, we have not resolved the discrepancies between our individual cohort data, in which we report greater sex differences in immunogenicity of seasonal A/H3N2 and pandemic A/H1N1 among reproductive-aged than older adults^3,6^, and other epidemiological analyses of hospitalization data for vaccine effectiveness (VE), which have reported more robust sex differences, with greater VE among females than males, in older adults (e.g., ≥65 years of age)^7,47^. These discrepancies may be due to significantly fewer incidences of severe disease and hospitalizations among reproductive aged individuals. Resolving these discrepancies is critical for to ensure that sex continues to be used as an effect modifier rather than a demographic variable in vaccine studies^2^.

Reproductive aged females can mount greater antibody responses than males to self (e.g., SLE) or foreign antigens (e.g., vaccines). Evolutionarily, females that develop higher antibody titers against diverse pathogens may have a reproductive advantage because this facilitated placental antibody transfer and provided protection of the offspring during the first months of life^48^. The heightened antibody responses to self-antigens by females might be an evolutionary side effect of changing environmental exposures, including to viruses^49^. Mechanistically, we^6,50,51^ and others^52^ have shown that both the sex steroid milieu (i.e., greater concentrations of estrogens) and the escape of *TLR7* from X chromosome inactivation play roles in increasing B cell production of antibodies against foreign and self-antigens. The next generation of vaccine clinical trials should consider sex as a biological variable as there is mounting evidence that this factor causes heterogeneity in vaccine outcomes.

## Methods

### Study design

100 healthcare workers (n=50/sex) at Johns Hopkins Health System (JHHS) ages 18-49 years old were recruited and enrolled in our study through the Johns Hopkins Centers of Excellence for Influenza Research and Response (JHCEIRR) annual vaccination program during fall of 2022-2023. Johns Hopkins Institutional Review Boards (IRB00091667) gave ethical approval for this work. Written consent was obtained for all participants during enrollment. Participant demographics were self-reported. Study data were collected and managed via REDCap electronic data capture tools hosted at Johns Hopkins University ^53,54^. After the pre-vaccination blood draw (0 DPV), standard dose inactivated quadrivalent influenza vaccines was administered intramuscularly. Blood samples were collected at 0-, 28-, and 180-days post-vaccination (DPV). Peripheral blood mononuclear cells (PBMCs) and plasma were separated and stored in liquid nitrogen or at −80°C, respectively.

### Influenza vaccine

The 2022-2023 quadrivalent influenza vaccine (QIV) formulation consisted of two influenza A and two influenza B strains based WHO recommendations for the Northern Hemisphere for the 2022-2023 season: A/Victoria/2570/2019 (H1N1) pdm09-like virus, A/Darwin/9/2021 (H3N2)-like virus, B/Austria/1359417/2021-like virus (Victoria lineage), and B/Phuket/3073/2013-like virus (Yamagata lineage).

### Viruses and recombinant HA antigens

The influenza A/Victoria/2570/2019 (H1N1) pdm09-like virus, influenza A/Darwin/9/2021 (H3N2)-like virus and B/Austria/1359417/2021–like virus were obtained from the US CDC. Responses to the B/Yamagata lineage were not measured because no naturally occurring B/Yamagata viruses have been confirmed since March 2020. The infectious virus titer was determined on MDCK-Siat cells using a 50% tissue culture infectious dose (TCID50) assay. For flow cytometry, glycosylated recombinant H3 protein with a C-terminal 8x His-tag (A/H3N2/Darwin/9/2021, eEnzyme # IA-H3-D21WP) was used. For flow cytometry staining, the recombinant H3 proteins were biotinylated using the EZ-Link Micro NHS-PEG4-Biotinylation Kit (Thermo Fisher #21455) per manufacturer’s instructions.

### Microneutralization

Plasma samples were diluted with receptor-destroying enzyme (RDE, Denka Seiken) at 1:3 ratio and incubated overnight at 37 °C followed by heat inactivation at 56 °C for 30 min. Samples were 2-fold serially diluted in Incomplete media (IM), composed of DMEM with 1%P/S and L-Glutamine (Incomplete medium, IM), mixed with 100 TCID50 of each virus A/Victoria/2570/2019 (H1N1) or A/Darwin/9/2021 (H3N2) or B/Austria/1359417/2021 (Victoria lineage), and incubated at room temperature for 1 h. The virus/plasma mixture was transferred in duplicate into the 96-well cell culture plates containing confluent SIAT1(cDNA of human 2,6-sialyltransferase) stable Expressing Madin-Darby Canine Kidney (MDCK-SIAT1) cells and incubated at 32 °C. After a 24 h incubation, plates were washed once with 1X PBS, fresh IM was added, and cells were incubated for 6 days. Plates were fixed with 4% formaldehyde, stained with naphthol blue-black solution, and scored as described previously^6,8^.

### Memory B cell ELISpot

IgG memory B cell responses against H3 were evaluated by ELISpot following B cell polyclonal stimulation and differentiation into antibody-secreting cells (ASCs)^55,56^. PBMCs were cultured for 7 days in phenol red-free RPMI 1640 medium (Gibco, #11835030) supplemented with 10% charcoal-stripped FBS (Gibco, #12676029 ), 2 mM L-glutamine (Gibco, # A2916801), 100 U/mL penicillin-streptomycin (Gibco, #15140-122), 10 mM HEPES (Gibco, #15630080), 2.5 mM sodium pyruvate (Sigma-Aldrich, #S8636), 1× essential amino acids (Gibco, # 11130051), and 1× nonessential amino acids (Gibco, #11140050). For polyclonal stimulation, PBMC were supplemented with 50 μM 2-mercaptoethanol (Bio-Rad, # 1610710), 6 μg/mL CpG oligodeoxynucleotide ODN-2006 (InvivoGen, #tlrl-2006), 1.2 μg/mL Protein A from *Staphylococcus aureus* Cowan strain (Sigma-Aldrich, #82526), and pokeweed mitogen (PWM; 1:100,000 dilution – a generous gift from Dr. S. Crotty). After 7 days of stimulation, the expanded cells were harvested, counted, and seeded in duplicate onto MultiScreen Cellulose Esters plates (Millipore-Sigma, #MAHVN4510) pre-coupled either with H3 from A/Darwin/9/2021-H3N2 (eEnzyme, # IA-H3-D21WP) or goat anti-human IgG antibody (Jackson ImmunoResearch, #109-005-098). Goat anti-human IgG was used to determine the total number of cells secreting IgG and served as a positive control. Negative controls involved wells coated with 1x PBS. Plates were incubated for 5 h at 37°C in 5% CO₂, washed, and incubated overnight with HRP-conjugated goat anti-human IgG (Jackson ImmunoResearch, #109-035-008, 1:1,000 dilution). Spots were developed using 3-amino-9-ethylcarbazole (AEC) substrate (BD Biosciences, #551951). Spot-forming cells (SFCs) and spot size were enumerated using an automated ELISpot reader (ImmunoSpot; Cellular Technology Limited, OH, USA) and analyzed using ImmunoSpot software version 7.0.26.0 Pro DC. Total and antigen-specific memory IgG B-cell responses were expressed as SFCs per 10⁶ cultured PBMCs.

### Luminex Antigen-Bead Coupling

H3 antigen (eEnzyme # IA-H3-D21WP) was covalently coupled to distinct Luminex MagPlex bead regions via carboxylate modification. Beads were activated using activation buffer (0.1 M NaH2PO4, pH 6.2) supplemented 25uL of both 50 mg/mL EDC and 50 mg/mL Sulfo-NHS. Activated beads were coupled to 25 µg of antigen in coupling buffer (0.05 M N-Morpholino ethanesulfonic acid, pH 5.0), blocked with PBS containing 0.1% BSA, 0.02% Tween-20, and 0.05% sodium azide (pH 7.4), and washed with PBS containing 0.05% Tween-20. Beads were stored at 4°C in PBS until use.

### Luminex-Based Isotyping and Fc Receptor Binding

Plasma samples (5 µL, diluted 1:25) were incubated with H3-coupled bead solution (20 µL per bead region) in Luminex Assay Buffer (PBS, 0.1% BSA, 0.05% Tween-20) for 2 hours on a plate shaker. Plates were placed on a magnet and washed with Procartaplex Wash Buffer (Thermo Fisher). Isotype/subclass detection was performed using PE-conjugated secondary antibodies (Southern Biotech) targeting IgM (# 9020-09, diluted 1:100), IgG (subclasses IgG1–4, (#9040-09, #9052-09, #9070-09, #9210-09, #9200-09, diluted 1:200), and IgA (subclasses IgA1–2 (#9130-09, #9140-09, diluted 1:100C-terminal polyhistidine tag and were biotinylated using the EZ-Link Micro NHS-PEG4-Biotinylation Kit and conjugated to PE-Streptavidin (#554061, BD biosciences)) for 15 minutes on a rotor prior to use. FcRs assessed included FcγRIIA-R, FcγRIIA-H FcγRIIB, FcγRIIIA-V, FcγRIIIA-F, FcγRIIIB, FcRn, and FcαR (Duke Human Vaccine Institute Protein Production Facility). Mean fluorescence intensity (MFI) was measured on a Luminex Intelliflex (FlexMap 3D instrument, low PMT; doublet discrimination range 7,000– 17,000; minimum 100 events per antigen). PBS served as a negative control, and positive controls were included in each run to ensure assay integrity.

### Antibody-Dependent Complement Deposition

Biotinylated H3-coupled beads were incubated with plasma samples (10uL, diluted 1:25) to form immune complexes, then washed three times with 1% BSA. Lyophilized Low-Tox Guinea Pig Complement (#CL4051, Cedarlane) was reconstituted in ice-cold dH2O, diluted 1:25 in GVB^++^ (Gelatin Veronal buffer with Mg2^+^ and Ca2^+^, #IBB-300, Boston Bio), and added to each well for incubation at 37°C. Complement deposition was detected using goat anti-guinea pig C3 IgG (1:500; #SKU0855385, MP Biomedicals) followed by donkey anti-goat IgG-PE secondary antibody (1:200; Thermo Fisher). Beads were resuspended and acquired on the Luminex Intelliflex as described above.

### Multiplexed Antibody-Dependent Non-Neutralizing Functional Flow Cytometry

A multiplex assay was performed to evaluate non-neutralizing antibody functions of H3-specific antibodies mediated through Fc receptor (FcR) activity across innate immune cell populations, including natural killer (NK) cells, monocytes, and neutrophils. Biotinylated antigen was generated using a 20-fold molar excess of biotin reagent and purified using spin columns to remove unbound biotin. The biotinylated antigen was subsequently coupled to fluorescent neutravidin beads by incubation overnight at 4°C, followed by washing and resuspension in PBS containing 0.1% BSA. Plasma samples were thawed and diluted 1:100 in PBS, then incubated with antigen-coupled beads for 2 hours at 37°C to allow immune complex formation. Whole blood–derived leukocytes were isolated from a female donor leukopak using ammonium chloride lysis, washed, and resuspended in RPMI supplemented with 10% fetal bovine serum and 1% penicillin-streptomycin (R10 Media). Cells were counted, adjusted to 1.5 × 10⁶ cells/mL, and stimulated overnight with IL-15 (1 ng/mL) at 37°C. Effector cells from this single female donor were used across all plasma samples, which were assayed together under identical conditions, to minimize inter-donor variability in Fc-mediated effector function. Following immune complex formation, beads were washed and incubated with effector cells at a density of 1 × 10⁵ cells per well. An inhibitor cocktail containing anti-CD107a and Golgi transport inhibitor was added, and plates were incubated for 5 hours at 37°C. Cells were then stained for extracellular markers to identify NK cells (CD3⁻CD56⁺CD66b⁻CD14⁻), neutrophils (CD3⁻CD66b⁺), and monocytes (CD3⁻CD66b⁻CD14⁺), as well as Fc receptors (CD64, CD32a and CD32b). Following surface staining, cells were fixed, permeabilized, and stained intracellularly for cytokines. NK cells engaged by the Fc region of H3-specific antibodies bound to the beads were identified as anti-H3-specific-NK cells (Figure S3). NK cell function was assessed as the frequency of anti-H3-specific NK cells positive for degranulation (CD107a), cytotoxic granule content (Granzyme B), and intracellular cytokine production (IFNγ and CCL4 [MIP-1β]). Phagocytic activity in neutrophils and monocytes was quantified based on bead uptake (bead⁺ cells). Beads incubated with effector cells in the absence of plasma served as negative controls. Monocytes were evaluated as total cells and within FcγR-defined subsets (CD64⁺, CD32a⁺, CD32b⁺), and neutrophils as total cells and within CD64⁺ and CD32a⁺ subsets. Samples were acquired on a BD FACSymphony A3 flow cytometer (BD Biosciences). PhagoScores were calculated as the percentage of bead⁺ cells multiplied by the geometric mean fluorescence intensity (GMFI) of bead⁺ cells, divided by 10,000 ^57^(**Figure S3**).

### Statistical Analyses

All neutralizing and systems serology data were log-transformed (log_2_ or log_10_) and non-neutralizing data generated by flow cytometry were arcsine-square root transformed to approximate Gaussian distribution. Differences in proportions (e.g., seroconversion and seroprotection rates) were compared by Fisher’s exact test. ELISpot count data were analyzed by Poisson mixed effects regression. Linear mixed effects models, adjusting for continuous age, were used to compare sex by time differences for serological data. Benjamini-Hochberg with an FDR of 0.05 was used to adjust for multiple comparisons. Sparse partial least squares discriminant analysis (sPLS-DA)^58^, a supervised machine learning algorithm with LASSO feature selection, was used to integrate all complete, non-missing systems serology and neutralizing antibody data from 0, 28, 180 DPV to classify sex. Mahalanobis distance was first used for principal component analysis (PCA) to visualize separation of females and males in the latent space. The number of components and features were tuned using Mahalanobis distance to establish the final model. 5-fold cross-validation with 50 repeats was used calculate an error rate and to evaluate model performance. Analyses and data visualizations were conducted in GraphPad Prism, Stata 17.0^59^, and RStudio^60^ 2026.7.1.14 using R4.5.3.

## Data Availability

The clinical data and specimens used in this study are from the Johns Hopkins Health System (JHHS). These individual-level patient data are protected for privacy. Qualified researchers affiliated with Johns Hopkins University (JHU) may apply for access through the Johns Hopkins Institutional Review Board (IRB; https://www.hopkinsmedicine.org/institutional_review_board/). Those not affiliated with JHU seeking to collaborate may contact the corresponding author. The datasets generated and/or analyzed during the current study are not publicly available due to IRB restrictions but are available from the corresponding author on reasonable request. Data in figures can be found at Vivli (https://doi.org/10.25934/PR00018381). All code can be accessed through GitHub at https://github.com/ayin0510/JHU85-2022-2023-Influneza-Vaccine-Responses-in-HCWs.

## Acknowledgments

We thank the clinical coordinators, nurses, and staff who assisted with sample collection. We thank the participants who volunteered to participate in this research. We thank the members of the Klein, Davis, and Thompson labs for detailed discussions about these data.

## Author contributions

Conceptualization: SLK, AP

Methodology: HSP, AY, FRT, SLK

Investigation: HSP, MAP, JH, AZ, RP

Visualization: HSP, AY, AZ

Funding acquisition: RER, AP, SLK

Project administration: KZJF, RER, AP, SLK

Supervision: KZKF, RER, FRT, SLK

Writing – original draft: HSP, AY, SLK

Writing – review & editing: all authors

## References

1 Shapiro, J. R. et al. Sex-specific effects of aging on humoral immune responses to repeated influenza vaccination in older adults. NPJ Vaccines 6, 147, doi:10.1038/s41541-021-00412-6 (2021).

2 Shapiro, J. R., Klein, S. L. & Morgan, R. Stop ’controlling’ for sex and gender in global health research. BMJ Glob Health 6, doi:10.1136/bmjgh-2021-005714 (2021).

3 Potluri, T. et al. Age-associated changes in the impact of sex steroids on influenza vaccine responses in males and females. NPJ Vaccines 4, 29, doi:10.1038/s41541-019-0124-6 (2019).

4 Furman, D. et al. Systems analysis of sex differences reveals an immunosuppressive role for testosterone in the response to influenza vaccination. Proc Natl Acad Sci U S A 111, 869–874, doi:10.1073/pnas.1321060111 (2014).

5 Engler, R. J. et al. Half-vs full-dose trivalent inactivated influenza vaccine (2004-2005): age, dose, and sex effects on immune responses. Arch Intern Med 168, 2405–2414, doi:168/22/2405 [pii]10.1001/archinternmed.2008.513 (2008).

6 Park, H. S. et al. Sex and age differences in antibody responses to seasonal influenza vaccination are mediated by estrogenic upregulation of NF-kappaB and TNF signaling in B cells. mBio, e0143526, doi:10.1128/mbio.01435-26 (2026).

7 Chambers, C. et al. Should Sex Be Considered an Effect Modifier in the Evaluation of Influenza Vaccine Effectiveness? Open Forum Infect Dis 5, ofy211, doi:10.1093/ofid/ofy211 (2018).

8 Ursin, R. L. et al. Differential antibody recognition of H3N2 vaccine and seasonal influenza virus strains based on age, vaccine status, and sex in the 2017-18 season. J Infect Dis, doi:10.1093/infdis/jiaa289 (2020).

9 Tadount, F. et al. Sex Differences in the Immunogenicity and Efficacy of Seasonal Influenza Vaccines: A Meta-analysis of Randomized Controlled Trials. Open Forum Infect Dis 11, ofae222, doi:10.1093/ofid/ofae222 (2024).

10 Boudreau, C. M. & Alter, G. Extra-Neutralizing FcR-Mediated Antibody Functions for a Universal Influenza Vaccine. Front Immunol 10, 440, doi:10.3389/fimmu.2019.00440 (2019).

11 Pyzik, M. et al. The Neonatal Fc Receptor (FcRn): A Misnomer? Front Immunol 10, 1540, doi:10.3389/fimmu.2019.01540 (2019).

12 DiLillo, D. J., Tan, G. S., Palese, P. & Ravetch, J. V. Broadly neutralizing hemagglutinin stalk-specific antibodies require FcgammaR interactions for protection against influenza virus in vivo. Nat Med 20, 143–151, doi:10.1038/nm.3443 (2014).

13 Leon, P. E. et al. Optimal activation of Fc-mediated effector functions by influenza virus hemagglutinin antibodies requires two points of contact. Proceedings of the National Academy of Sciences of the United States of America 113, E5944–E5951, doi:10.1073/pnas.1613225113 (2016).

14 Vanderven, H. A. C Kent, S. J. The protective potential of Fc-mediated antibody functions against influenza virus and other viral pathogens. Immunol Cell Biol 98, 253–263, doi:10.1111/imcb.12312 (2020).

15 Henry Dunand, C. J., et al. Both Neutralizing and Non-Neutralizing Human H7N9 Influenza Vaccine-Induced Monoclonal Antibodies Confer Protection. Cell host & microbe 19, 800–813, doi:10.1016/j.chom.2016.05.014 (2016).

16 Ermler, M. E. et al. Chimeric Hemagglutinin Constructs Induce Broad Protection against Influenza B Virus Challenge in the Mouse Model. J Virol 91, doi:10.1128/JVI.00286-17 (2017).

17 McMahon, M. et al. Vaccination With Viral Vectors Expressing Chimeric Hemagglutinin, NP and M1 Antigens Protects Ferrets Against Influenza Virus Challenge. Front Immunol 10, 2005, doi:10.3389/fimmu.2019.02005 (2019).

18 Broecker, F. et al. A mosaic hemagglutinin-based influenza virus vaccine candidate protects mice from challenge with divergent H3N2 strains. NPJ Vaccines 4, 31, doi:10.1038/s41541-019-0126-4 (2019).

19 Nachbagauer, R. et al. A chimeric hemagglutinin-based universal influenza virus vaccine approach induces broad and long-lasting immunity in a randomized, placebo-controlled phase I trial. Nat Med 27, 106–114, doi:10.1038/s41591-020-1118-7 (2021).

20 Nachbagauer, R. et al. Age Dependence and Isotype Specificity of Influenza Virus Hemagglutinin Stalk-Reactive Antibodies in Humans. mBio 7, e01996–01915, doi:10.1128/mBio.01996-15 (2016).

21 Hansen, C. S. et al. Sex shapes CD64 expression and vaccine-induced monocytic responses. Biol Sex Differ 17, doi:10.1186/s13293-026-00897-7 (2026).

22 Miller-Novak, L. K. et al. Analysis of Complement-Mediated Lysis of Simian Immunodeficiency Virus (SIV) and SIV-Infected Cells Reveals Sex Differences in Vaccine-Induced Immune Responses in Rhesus Macaques. J Virol 92, doi:10.1128/JVI.00721-18 (2018).

23 Chaulagain, S., Liu, J. A., McCombs, J. E. & Klein, S. L. Sex differences in immune responses to viruses, bacteria and vaccines. Nat Immunol 27, 660–673, doi:10.1038/s41590-026-02470-1 (2026).

24 Boudreau, C. M. et al. Pre-existing Fc profiles shape the evolution of neutralizing antibody breadth following influenza vaccination. Cell Rep Med 4, 100975, doi:10.1016/j.xcrm.2023.100975 (2023).

25 Bruhns, P. et al. Specificity and affinity of human Fcγ receptors and their polymorphic variants for human IgG subclasses. Blood 113, 3716–3725, doi:10.1182/blood-2008-09-179754 (2009).

26 Marchese, A. M. et al. Mechanisms and implications of IgG4 responses to SARS-CoV-2 and other repeatedly administered vaccines. Journal of Infection 89, 106317, doi:10.1016/j.jinf.2024.106317 (2024).

27 Bournazos, S. C Ravetch, J. V. Fcgamma Receptor Function and the Design of Vaccination Strategies. Immunity 47, 224–233, doi:10.1016/j.immuni.2017.07.009 (2017).

28 Tzeng, S. J., Li, W. Y. & Wang, H. Y. FcgammaRIIB mediates antigen-independent inhibition on human B lymphocytes through Btk and p38 MAPK. J Biomed Sci 22, 87, doi:10.1186/s12929-015-0200-9 (2015).

29 Tang, C. C. et al. Phenotypical and functional profiles of natural killer cells exhibiting matrix metalloproteinase-mediated CD16 cleavage after anti-HIV antibody-dependent activation. Clin Exp Immunol 181, 275–285, doi:10.1111/cei.12593 (2015).

30 Romee, R. et al. NK cell CD16 surface expression and function is regulated by a disintegrin and metalloprotease-17 (ADAM17). Blood 121, 3599–3608, doi:10.1182/blood-2012-04-425397 (2013).

31 Wang, Y. et al. ADAM17 cleaves CD16b (FcgammaRIIIb) in human neutrophils. Biochim Biophys Acta 1833, 680–685, doi:10.1016/j.bbamcr.2012.11.027 (2013).

32 Jing, Y. et al. Identification of an ADAM17 cleavage region in human CD16 (FcgammaRIII) and the engineering of a non-cleavable version of the receptor in NK cells. PLoS One 10, e0121788, doi:10.1371/journal.pone.0121788 (2015).

33 Marzeda, A. M. et al. Investigating endogenous immune-mediated monocyte memory in rheumatoid arthritis. Ann Rheum Dis 84, 1484–1500, doi:10.1016/j.ard.2025.03.016 (2025).

34 Karsten, C. B. et al. A versatile high-throughput assay to characterize antibody-mediated neutrophil phagocytosis. J Immunol Methods 471, 46–56, doi:10.1016/j.jim.2019.05.006 (2019).

35 Zohar, T. et al. A multifaceted high-throughput assay for probing antigen-specific antibody-mediated primary monocyte phagocytosis and downstream functions. J Immunol Methods 510, 113328, doi:10.1016/j.jim.2022.113328 (2022).

36 Centers for Disease, C. & Prevention. Influenza Activity in the United States during the 2022–2023 Season and Composition of the 2023–2024 Influenza Vaccine. (2023).

37 Merced-Morales, A. et al. Influenza Activity and Composition of the 2022–23 Influenza Vaccine — United States, 2021–22 Season. MMWR Morb Mortal Wkly Rep 71, 913–919, doi:10.15585/mmwr.mm7129a1 (2022).

38. (!!! INVALID CITATION !!! 6,41).

39 Richardson, S. I. et al. IgG3 enhances neutralization potency and Fc effector function of an HIV V2-specific broadly neutralizing antibody. PLOS Pathogens 15, e1008064, doi:10.1371/journal.ppat.1008064 (2019).

40 Richardson, S. I. et al. HIV Broadly Neutralizing Antibodies Expressed as IgG3 Preserve Neutralization Potency and Show Improved Fc Effector Function. Frontiers in Immunology 12, 733958, doi:10.3389/fimmu.2021.733958 (2021).

41. (!!! INVALID CITATION !!! ).

42 Lin, G. G. C Li, J. M. IgG subclass serum levels in systemic lupus erythematosus patients. Clin Rheumatol 28, 1315–1318, doi:10.1007/s10067-009-1224-x (2009).

43 Pan, Ǫ., et al. IgG4 Autoantibodies Attenuate Systemic Lupus Erythematosus Progression by Suppressing Complement Consumption and Inflammatory Cytokine Production. Front Immunol 11, 1047, doi:10.3389/fimmu.2020.01047 (2020).

44. 44 Anaya, E. U. et al. Antibody Transcytosis and Neutralizing Activity in Respiratory Epithelial Cells. *bioRxiv*, doi:10.64898/2026.05.25.727697 (2026).

45 Anthi, A. K. et al. An intranasal subunit vaccine induces protective systemic and mucosal antibody immunity against respiratory viruses in mouse models. Nat Commun 16, 3999, doi:10.1038/s41467-025-59353-6 (2025).

46 Ochsner, S. P. et al. FcRn-Targeted Mucosal Vaccination against Influenza Virus Infection. J Immunol 207, 1310–1321, doi:10.4049/jimmunol.2100297 (2021).

47 Harker, E. J. et al. Differences in influenza vaccine effectiveness by sex among adults hospitalized with acute respiratory illness-IVY network, January 24, 2022-September 1, 2024. Vaccine 74, 128192, doi:10.1016/j.vaccine.2025.128192 (2026).

48 Fink, A. L. C Klein, S. L. The evolution of greater humoral immunity in females than males: implications for vaccine efficacy. Current Opinion in Physiology 5, 16–20 (2018).

49 Barrie, W. et al. Elevated genetic risk for multiple sclerosis emerged in steppe pastoralist populations. Nature 625, 321–328, doi:10.1038/s41586-023-06618-z (2024).

50 Fink, A. L., Engle, K., Ursin, R. L., Tang, W. Y. & Klein, S. L. Biological sex affects vaccine efficacy and protection against influenza in mice. Proc Natl Acad Sci U S A 115, 12477–12482, doi:10.1073/pnas.1805268115 (2018).

51 Dhakal, S. et al. Estradiol mediates greater germinal center responses to influenza vaccination in female than male mice. mBio 15, e0032624, doi:10.1128/mbio.00326-24 (2024).

52 Souyris, M. et al. TLR7 escapes X chromosome inactivation in immune cells. Science Immunology 3, eaap8855, doi:10.1126/sciimmunol.aap8855 (2018).

53 Harris, P. A. et al. Research electronic data capture (REDCap)--a metadata-driven methodology and workflow process for providing translational research informatics support. J Biomed Inform 42, 377–381, doi:10.1016/j.jbi.2008.08.010 (2009).

54 Harris, P. A. et al. The REDCap consortium: Building an international community of software platform partners. J Biomed Inform 95, 103208, doi:10.1016/j.jbi.2019.103208 (2019).

55 Toapanta, F. R. et al. Further characterization of Shigella-specific (memory) B cells induced in healthy volunteer recipients of SF2a-TT15, a Shigella flexneri 2a synthetic glycan-based vaccine candidate. Front Immunol 14, 1291664, doi:10.3389/fimmu.2023.1291664 (2023).

56 Crotty, S., Aubert, R. D., Glidewell, J. & Ahmed, R. Tracking human antigen-specific memory B cells: a sensitive and generalized ELISPOT system. J Immunol Methods 286, 111–122, doi:10.1016/j.jim.2003.12.015 (2004).

57 Lu, L. L. et al. IFN-gamma-independent immune markers of Mycobacterium tuberculosis exposure. Nat Med 25, 977–987, doi:10.1038/s41591-019-0441-3 (2019).

58 Rohart, F., Gautier, B., Singh, A. & Le Cao, K. A. mixOmics: An R package for ’omics feature selection and multiple data integration. PLoS Comput Biol 13, e1005752, doi:10.1371/journal.pcbi.1005752 (2017).

59. Stata Statistical Software: Release 17 v. Release 17 (StataCorp LLC, College Station, TX, 2025).

60. RStudio: Integrated Development Environment for R (Posit Software, PBC, Boston, MA, 2026).

