## Supplementary Materials for "Broader functional and memory antibody responses in adult females than males in response to seasonal H3N2 vaccine virus"

**Figure S1: Seroprotection, seroconversion, and titers against A/H1N1 and B/Victoria.** Virus neutralizing antibody (nAb) titers against (**a**) A/H1N1 and (**b**) B/Victoria were measured from serum collected at 0, 28, and 180 DPV. Data were $log_2$-transformed and analyzed based on seroconversion status for each virus (responder: $\geq$4 FC; non-responder: <4FC) using linear mixed-effects regression models, adjusting for continuous age. † represents $p<0.05$ timepoint differences. Brackets for timepoint differences are colored by sex (green=males, purple=females). Proportion of individuals that reached seroprotective (SP; 1:40 titer) antibody levels are shown. The limit of detection (LOD) for the assay was 1:10 titer or 3.32 $log_2$(titer).

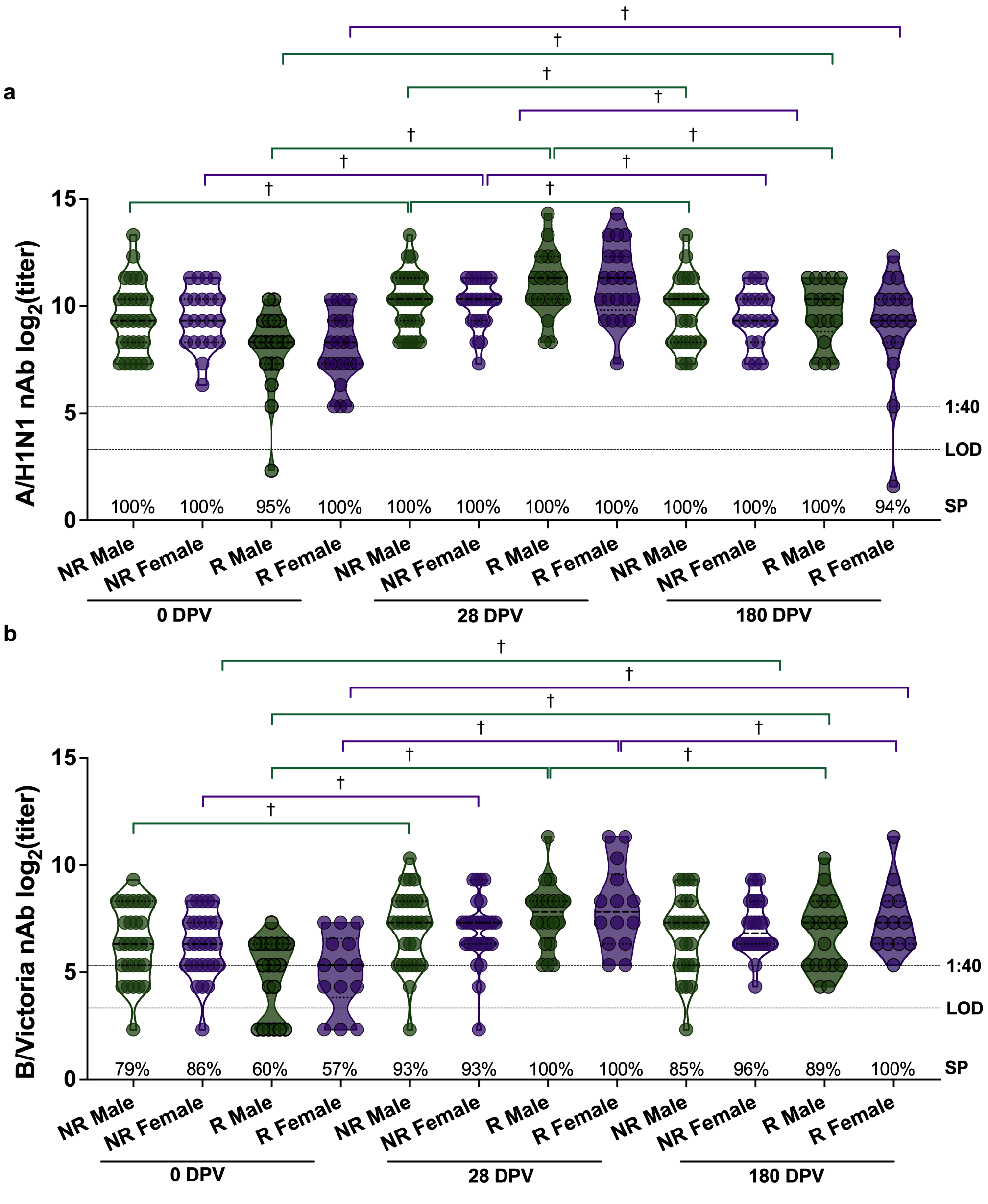

**Figure S2: Antibody-dependent complement deposition (ADCD) and neutrophil phagocytosis (ADNP) in H3N2 responders.** ADCD was analyzed from plasma with a Luminex-based assay and ADNP was assessed using effector neutrophils obtained from donor leukocytes by flow cytometry. Plasma from H3N2 responders collected at 0, 28, and 180 DPV was used. (**a**) ADCD mean fluorescence intensity (MFI) of antibodies binding C3 and ADNP PhagoScores for total (**b**), CD64$^+$ (**c**) and CD32a$^+$ (**d**) neutrophils were $\log_{10}$-transformed and analyzed with linear mixed-effects regression modeling for sex and time differences, adjusting for continuous age. * represents $p<0.05$ sex differences and † represents $p<0.05$ timepoint differences. Brackets for timepoint differences are colored by sex (green=males, purple=females).

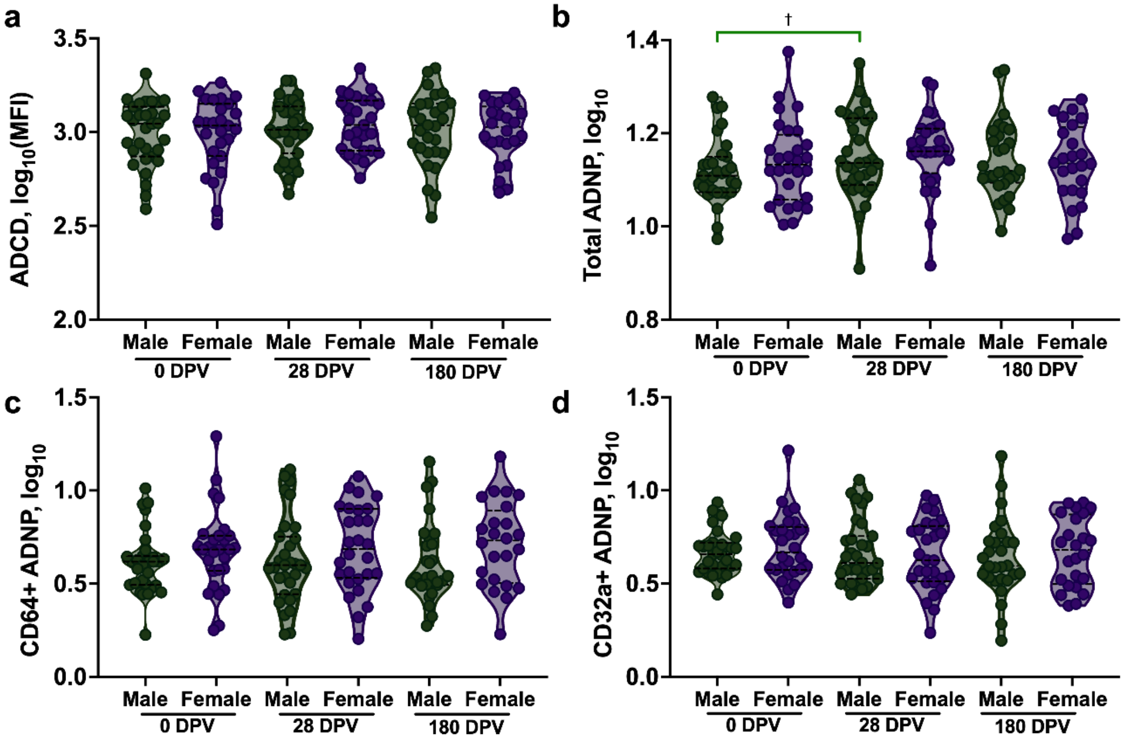

### Figure S3: Flow cytometry gating strategies for non-neutralizing functional analyses

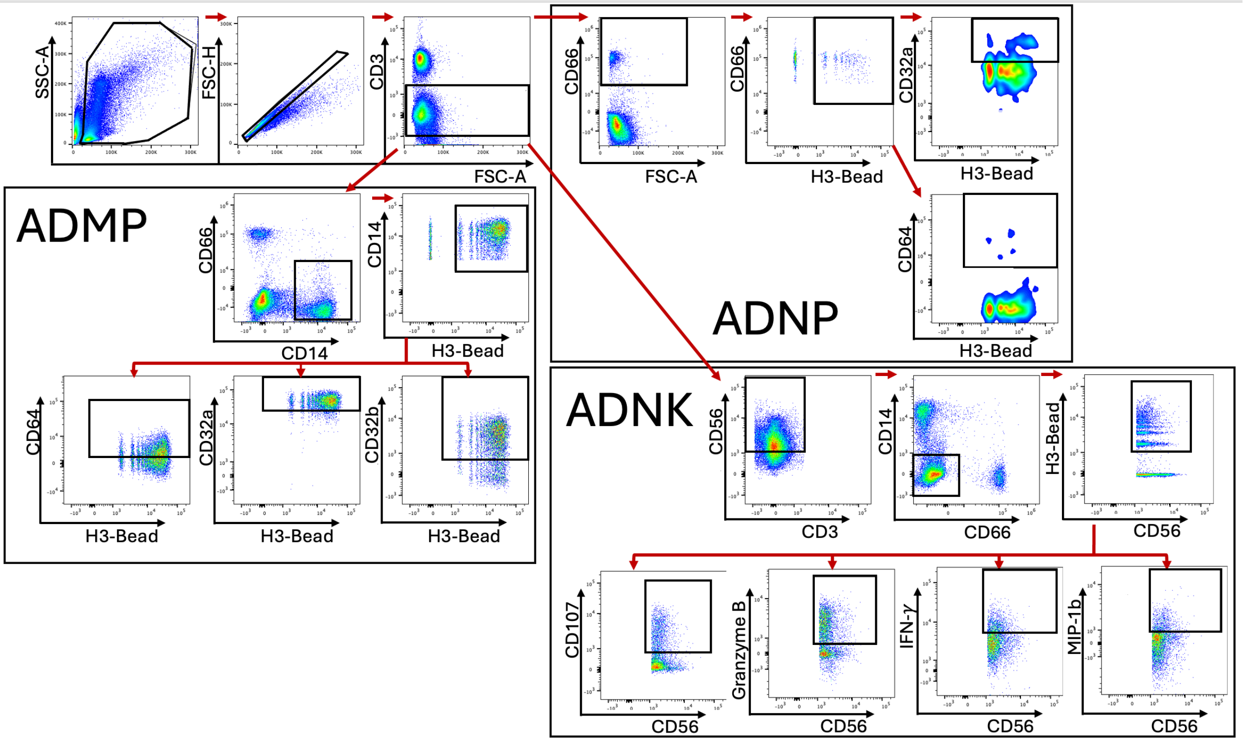

**Supplemental Table 1: Antibodies used for flow cytometry**

| Ab | Color | Cat # | vendor |
|---|---|---|---|
| CD32B | APC | 398304 | Biolegend |
| CD64 | BV605 | 305034 | Biolegend |
| CD3 | BV650 | 344872 | Biolegend |
| CD66B | PB | 305112 | Biolegend |
| CD32A | PE | 365504 | Biolegend |
| Granzyme B | Percp-cy5.5 | 372212 | Biolegend |
| CCL4 (MIP-1β) | Alexa700 | 561278 | BD |
| IFN-γ | BV510 | 563287 | BD |
| CD56 | PE-Cy7 | 557747 | BD |
| CD107a | PE-Cy5 | 555802 | BD |
| CD14 | BV786 | 563698 | BD |
